# Scalable Identification of Clinically Relevant COPD Documents: A Lightweight NLP Model for Large-Scale EHR Datasets

**DOI:** 10.1101/2025.04.22.25326240

**Authors:** Mohammed Al-Garadi, Sharon E. Davis, Michael E. Matheny, Dax Westerman, Adrienne K. Conger, Bradley W. Richmond, Thomas A. Lasko, Iben M. Ricket, Laura M. Paulin, Jeremiah R. Brown, Ruth M. Reeves

**Affiliations:** Department of Biomedical Informatics Vanderbilt University Medical Center Nashville TN; Geriatric Research Education and Clinical Care Center Tennessee Valley Healthcare System VA Nashville TN; Department of Biostatistics Vanderbilt University Medical Center Nashville TN; Division of General Internal Medicine Vanderbilt University Medical Center Nashville TN; Division of Allergy, Pulmonology, and Critical Care Medicine Vanderbilt University Medical Center Nashville TN; Department of Veterans Affairs Medical Center, Tennessee Valley Healthcare System Nashville TN; Department of Computer Science Vanderbilt University Nashville TN; Department of Epidemiology Dartmouth Geisel School of Medicine Lebanon, NH; Department of Pulmonology Dartmouth Hitchcock Medical Center Lebanon NH; Dartmouth Center for Implementation Science Geisel School of Medicine Lebanon, NH

## Abstract

**Background:** The widespread adoption of electronic health records (EHRs) has resulted in the generation of large volumes of clinical notes. Learning algorithms and large language models (LLMs) train from these resources but are susceptible to noise—irrelevant or non-informative data from them. This sensitivity can lead to significant challenges, including performance degradation and the generation of inaccurate predictions or “hallucinations.” This study addresses a critical challenge in clinical informatics: efficiently filtering millions of documents for relevance before advanced language model processing, particularly in resource-constrained environments. We present a novel framework for determining document relevance in clinical settings, utilizing a chronic obstructive pulmonary disease (COPD) dataset.

**Methods:** We developed a novel framework using weak supervision and domain-expert heuristics to generate “silver standard” labels for training data and expert annotated labels (gold stand),creating two datasets to optimize the model during the development phase and subsequent testing phase. Various text representation techniques, including Bag-of-Words, TF-IDF, lightweight document embeddings, compression-based features, and UMLS concept extraction, were evaluated. These representations were used to train Random Forest, XGBoost, and K-Nearest Neighbors classifiers. Models were optimized on a small expert-annotated dataset and evaluated on a held-out test set.

**Results:** The combination of lightweight document embedding with a Random Forest classifier demonstrated the best performance, achieving a precision of 0.75, recall of 0.89, and F1-score of 0.81 (95% CI: 0.76-0.87) for identifying relevant COPD documents. This significantly outperformed baseline heuristics (precision: 0.70, recall: 0.38, F1-score: 0.50, 95% CI: 0.43-0.56) and other tested methods.

**Conclusion:** Our study presents a novel framework for identifying COPD-relevant clinical documents using lightweight embedding and machine learning. This approach effectively filters pertinent documents, enhancing information retrieval precision. The framework’s scalability and minimal annotation needs make it promising for diverse healthcare applications, potentially optimizing clinical outcomes through efficient document selection for data-driven decision support systems.

## Background and related work

Chronic obstructive pulmonary disease (COPD) is a leading cause of mortality in the United States, impacting an estimated 24 million people nationwide^1^. COPD exacerbation-related hospitalizations also incur substantial healthcare costs, ranging from $7,000 to $39,200 per hospitalization^2^.

Overall, COPD-attributable care amounted to $49 billion in 2020, with hospitalization comprising approximately half of this total cost^3^.

Artificial intelligence (AI) is transforming healthcare by enhancing the analysis, interpretation, and prediction of medical data. This technological advancement supports clinical decision-making and enables more efficient allocation of limited resources for chronic disease management, particularly for high-risk patients^4 5^. While narrative clinical text is information-rich, its unstructured nature poses challenges for direct use in AI-driven clinical decision support systems.

To address this, information extraction and representation methods have emerged, leveraging large language models (LLMs) and deep learning techniques ^6–10^. These approaches aim to transform raw clinical narratives into structured, machine-readable data. However, significant challenges persist when processing vast document collections. Information relevant to a specific task may be dispersed across only a small fraction of the total corpus, making it inefficient to apply computationally intense LLMs to the entire dataset^11^. Sophisticated pre-processing, filtering, and prioritization algorithms can identify and extract relevant documents, reducing the volume of text that needs to be processed by computationally intense stages of the pipeline^12^.

### A. Weak supervision for developing automated NLP classification models

Weak supervision is a machine learning approach that mitigates the challenges of creating large, annotated datasets for NLP. It involves using lower-accuracy or less detailed programmatically generated labels and limited labeled data in semi-supervised learning^13–15^. This approach reduces the need for extensive manual labeling, speeding up the machine learning process and cutting costs. This method offers several advantages, including reduced annotation costs, improved scalability, and enhanced domain adaptability^13–15^. Weak supervision allows for the incorporation of domain expertise, supports iterative refinement, and enables model development in low-resource scenarios^13–15^. By minimizing manual labeling efforts, it facilitates the efficient creation of large-scale, domain-specific datasets, making it particularly valuable for developing robust NLP models across diverse fields ^16–18^.

Several recent studies have demonstrated the effectiveness of weak supervision in medical NLP tasks. Wang et al. combined this approach with pre-trained word embeddings to create a rule-based NLP algorithm for the automatic generation of training labels ^19^. They evaluated the effectiveness of these auto-generated labels across four supervised learning architectures: support vector machines, random forest (RF), fully connected networks, and convolutional neural networks. Their comparative analysis demonstrated the versatility and efficacy of this auto-labeling technique, as the generated labels performed successfully across diverse model frameworks ^19^.

Similarly, Cusick et al. (2021) applied weak supervision to address the challenge of exhaustive manual labeling in clinical contexts^18^. Building on a previous study that used a rule-based NLP system to automatically label clinical notes of 600 patients with potential suicidal ideation ^18^, they efficiently generated a sizable training dataset. This dataset was then used to train various statistical machine-learning models and a convolutional neural network. Their research highlighted the potential of combining weak supervision and deep learning to enhance real-time clinical systems and facilitate research on suicidal ideation progression through automated analysis of clinical text ^18^. Another study in this field came with the introduction of Trove by Fries et al. ^16^.

This framework for weakly supervised medical entity classification leverages existing medical ontologies like UMLS as a source of reusable, automated labeling heuristics. Trove’s key innovation is its use of a label model to learn the accuracies of individual ontologies and correct for label noise when combining multiple sources. The authors demonstrated weakly supervised performance on NER tasks for chemicals, diseases, and drugs, achieving results within 1.3-4.9 F1 points of fully supervised models.

These studies underscore the growing importance of weak supervision in overcoming the limitations of traditional manual annotation methods in clinical NLP tasks. By enabling the efficient creation of large-scale, domain-specific datasets, weak supervision is proving to be a valuable approach for developing robust NLP models across diverse clinical applications. This method offers a simplified way to auto-generate training labels, significantly reducing the need for manual annotation^16–19^.

Our proposed method uniquely integrates high-accuracy expert labels with weak supervision, offering a more efficient and accurate clinical document classification solution that overcomes traditional techniques’ limitations. In the following sections, we will describe the rationale, significance, and unique contributions of our proposed approach, demonstrating how it advances clinical NLP methods and addresses critical needs in the field.

### Rationale, importance, and contributions

NLP models designed for categorizing clinical documents can support various downstream AI/ML tasks in healthcare, such as risk prediction, clinical summarization, and probabilistic phenotyping ^20–22^. The recent introduction of LLMs has created tremendous potential for healthcare to leverage these powerful tools.

However, the bottleneck lies in the vast number of documents that these computationally intensive models need to process, which can reduce their efficiency and effectiveness. Lightweight NLP models solve this challenge by efficiently filtering documents and providing appropriate data inputs for more sophisticated models, such as LLMs ^10,23,24 25^. These lightweight models could potentially identify and select relevant documents, which may enhance the accuracy and efficiency of the overall AI/ML pipeline. However, the effectiveness of these models depends on input data quality. ICD codes, traditionally used to identify relevant documents, have proven ineffective ^26,27^. This observation underscores the limitations of relying exclusively on ICD codes for document selection in this context. The findings suggest the necessity for more refined and thorough identification methods to ensure the accurate capture of relevant COPD-related documents.

The potential effectiveness of these lightweight NLP models could stem from their ability to accurately identify and filter out irrelevant documents while potentially maintaining efficiency in computing power, memory usage, and processing time. This efficiency is especially advantageous for resource-constrained healthcare systems, where simpler machine learning approaches that require significantly less computational power are often preferred over advanced NLP techniques ^28^ such as LLMs.

By leveraging heuristic training data and a small expert-annotated dataset, these models can be optimized with minimal labeled data, significantly reducing the manual annotation burden. This approach addresses the persistent challenge of limited annotated data in clinical settings, potentially accelerating the development and deployment of AI-driven healthcare solutions.^19,29^

#### Contributions

Our study makes three primary contributions to the field of clinical document classification:

- We introduce a novel framework that uniquely integrates high-accuracy expert labels with weak supervision for hyperparameter tuning, thus designing lightweight machine learning classifiers. This approach offers a more efficient and accurate solution for categorizing COPD-related documents as either ‘relevant’ or ‘non-relevant,’ overcoming the limitations of traditional techniques.
- We leverage heuristic training data formulated by domain-expert clinicians to construct a training dataset without the need for extensive manual annotation. By using these expert hypotheses to generate “silver standard” labels^30,31^, we substantially mitigate the challenges commonly associated with manual annotation.
- We conduct extensive experiments with diverse text representation techniques, rigorously evaluating the relative performance of different pipelines for identifying relevant documents while minimizing the inclusion of irrelevant ones.

Although our study focuses on patients with COPD, the techniques we develop are designed to be adaptable across various clinical domains. Our approach of combining a small number of high-accuracy expert labels for hyperparameter tuning with a larger set of lower-accuracy labels for model training allows for potential improvements in model performance that may not be achievable using only lower-accuracy labels.

## Materials and Methods

We constructed a retrospective observational cohort of patients with COPD attending Vanderbilt University Medical Center (VUMC) between 01/01/2012 and 12/31/2020. A series of NLP processing pipelines of all clinical narrative notes from this cohort were used to perform data evaluation. The objective of the study was to create an AI/ML pipeline using weak supervision with a small set of annotated data that could effectively determine relevance of the notes for future AI/ML tasks for a clinical condition of interest, COPD. Our cohort inclusion criteria were patients with a diagnosis of COPD at any point during the study period and who were age 40 or greater at the time of their diagnosis. COPD was defined based on administrative codes available from the EHR for ICD-9-CM: 491, 491.1, 491.2, 491.21, 491.22, 491.8, 491.9, 492, 492.8, 493.92, 496 or ICD-10-CM: J41.0, J41.1, J41.8, J42, J43.1, J43.2, J43.8, J43.9, J44.0, J44.1, J44.9. This study was approved by the VUMC Institutional Review Board.

Figure 1 illustrates our NLP framework for classifying clinical notes related to COPD, including heuristic data creation, text representation, model training, optimization, and evaluation. Our heuristic training data, or “silver standard,” consists of 20,000 clinical notes. We created this dataset using expert-defined rules: notes from encounters with COPD diagnosis codes were labeled as informative (positive), while those from encounters without COPD codes or from 24+ months prior to initial COPD diagnosis, were labeled non-informative (negative). This heuristic approach enables the creation of a large training dataset without manual annotation, leveraging domain expertise to generate plausible labels. We employ various text representation techniques to transform the free text into numerical features, including Bag-of-Words (BoW), Term Frequency-Inverse Document Frequency (TF-IDF), Lightweight Document Embedding, Compression-based Embeddings, and UMLS (Unified Medical Language System) concept extraction. These representations feed into three machine learning classifiers: RF^32^, K-Nearest Neighbors (KNN)^33^, and Extreme Gradient Boosting (XGBoost)^34^. For model optimization and evaluation, we use a separate gold standard dataset manually annotated by clinical experts. We use 30% of this set for hyperparameter optimization, allowing us to fine-tune our models based on high-quality, expert-labeled data, and 70% as a held-out test set for final model assessment. By leveraging a large heuristically labeled dataset for training, and then optimizing and testing with a carefully curated gold standard, this approach integrates heuristic-based training data creation, diverse text representation techniques, and a two-stage evaluation process using expert-annotated data. The following subsections will explain each block in more detail.

**Figure 1.**
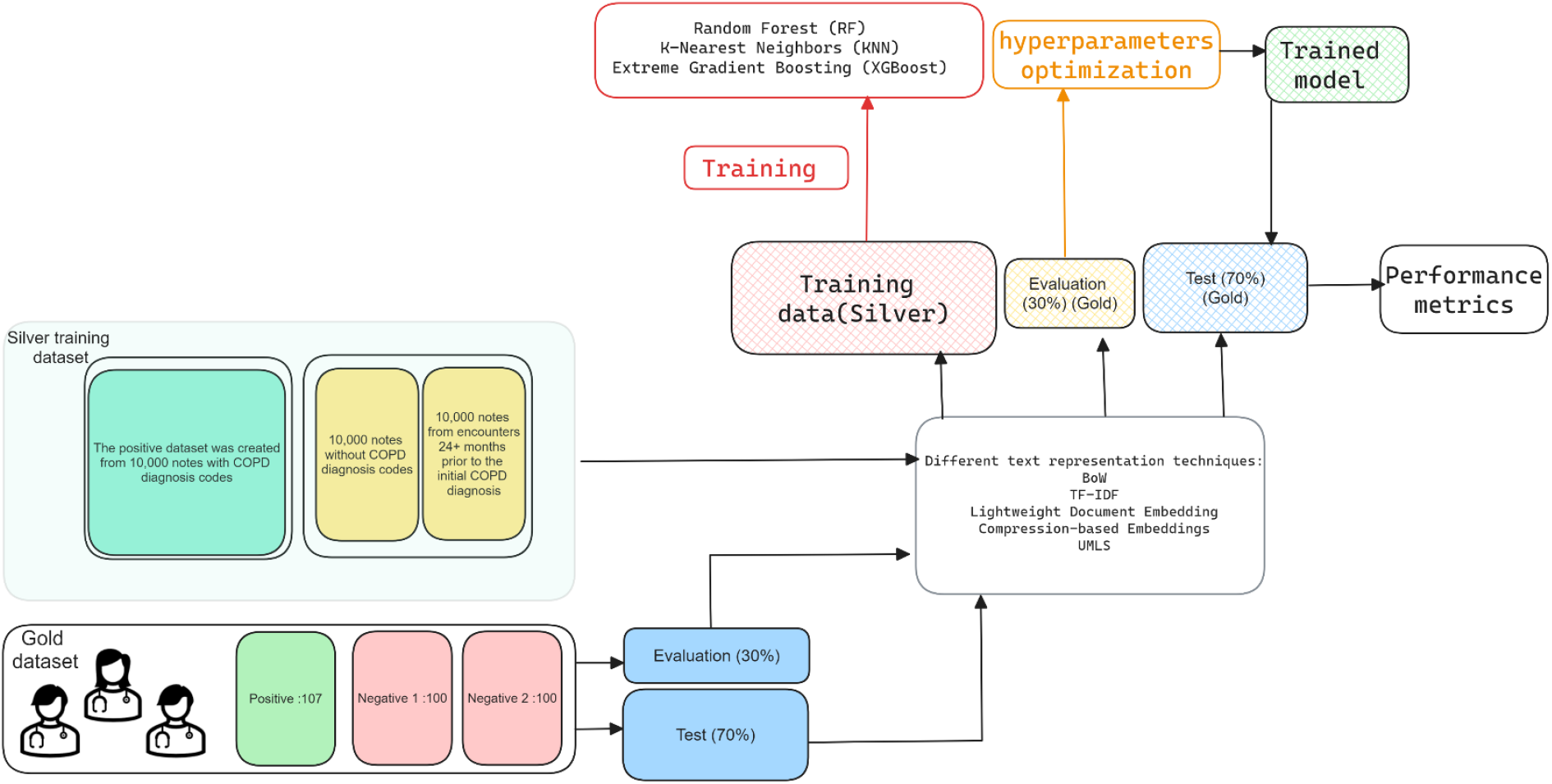
Framework for developing an effective lightweight classifier for clinical documents.

### 1. Data Collection and Annotation

This subsection details our data collection and labeling methods for identifying COPD-relevant clinical notes. We utilized a two-tiered strategy to build our training and test datasets: heuristic-based labeling for large-scale training (silver standard) and manual expert annotation for precise model optimization and testing (gold standard). This method effectively addresses the challenge in medical NLP of creating large, reliable training data while ensuring thorough model evaluation.

We compiled a corpus of 10 million clinical notes extracted from 32 million raw notes collected over a decade (study period). From this collection, we developed our training and test datasets:

#### Heuristic Training Dataset (Silver Standard)

The training dataset consists of positive and negative examples, identified using ICD code heuristics as follows:

A. COPD Encounter Corpus (Positive Labels): 10,000 notes from encounters ICD-coded as COPD, likely containing COPD-relevant information.
B. Non-COPD Encounter Corpus (Negative Labels): 10,000 notes from patient encounters without any COPD diagnosis code, indicating content likely irrelevant to COPD.
C. Pre-COPD Encounter Corpus (Negative Labels): 10,000 notes from patient encounters occurring 24+ months prior to the initial COPD diagnosis. This approach aims to avoid including notes with early signs of COPD

We constructed three training subsets to evaluate different heuristic combinations and their impact on classifier accuracy. Each subset contained 5,000 positive examples from A and 5,000 negative examples. The negative examples varied across subsets: the first used a mix of 2,500 each from B and C, the second used 5,000 from B only, and the third used 5,000 from C only.

This design allows us to assess which combination produces the most accurate classifier. We ensured diversity in patient demographics, timing, and note types across all samples.

#### Test data (Gold Standard)

Clinical experts developed a guideline to determine the relevance of documents for COPD diagnosis. This guideline was iteratively refined through consultations with clinical experts who provided valuable feedback. We then created a reference standard for evaluation and test sets by randomly selecting 307 documents from the positive (107) and negative training datasets (100 non-COPD and 100 Pre-COPD Encounter). Three clinicians independently reviewed and annotated these documents, labeling them as either relevant or irrelevant for COPD exacerbation prediction. The annotated documents were then divided, with 30% (92 documents) used for validation and model optimization during development and 70% (215 documents) held out for final testing.

### 2. NLP Model Development

The development of the NLP pipeline for classifying clinical documents related to COPD diagnosis involved several key stages. Initially, various NLP techniques were applied to transform clinical text into informative numerical representations. These numerical representations served as input for machine learning models, which were subsequently optimized to enhance performance.

Finally, the fully optimized models were assessed against held-out test set to measure their effectiveness. The following subsection will explain in detail the different text representation methods, machine learning approaches, and the processes of training, optimization, and testing the models.

#### Text representation

This step involves exploring various text representation methods, including Bag of Words, TF-IDF (Term Frequency-Inverse Document Frequency), Lightweight Document Embedding, and lossless compression representation.

1. BoW is a text representation technique that treats each document as a collection of words, disregarding their order. It creates a vector where each element corresponds to the frequency of a word in the document, ignoring context^35^.
2. TF-IDF is a text analysis method that evaluates the importance of words in a document within a larger corpus. It calculates a score for each word based on its frequency in the document (TF) and its rarity across all documents (IDF), helping to identify key terms^36^.
3. Lightweight Document Embedding ^37 38^. It is a modern text representation approach that uses pre-trained language models like Word2Vec or FastText to convert words or phrases into dense, continuous-valued vectors. These embeddings capture semantic relationships between words and are useful for various natural language processing tasks. Utilizing distributed memory and distributed bag of words approaches for enhancing paragraph and document embeddings ^38^, we incorporated the hierarchical softmax algorithm ^39^.
4. Compression representation. This approach utilizes lossless compression techniques. The key idea is to leverage compressors to efficiently represent text information, as they excel at capturing regularity. Normalized Compression Distance (NCD)^40 41^ quantifies shared information between text objects by comparing their compressed lengths, making it suitable for classification. Using gzip as the compressor, this lightweight and universal method calculates NCD distances between test and training data for KNN classification, offering a simple and resource-efficient alternative to deep neural networks for text classification tasks. This lightweight methodology is readily applicable to extensive text and has demonstrated better performance compared to the BERT model^41^. While this approach has been tested on general texts, such as news articles and Yahoo Answers, we extended its use by applying it to clinical documents. However, machine learning classifiers like RF and XGBoost do not natively utilize compression distance metrics as features. We thus transformed raw text via compression into the GZip format. The length of the compressed data then served as the key predictive feature for RF and XGBoost models.
5. UMLS text representation is a comprehensive resource developed by the National Library of Medicine (NLM) to facilitate the integration of biomedical information systems^42,43^. One of its key components is the representation of biomedical concepts and their relationships through various text formats. In UMLS, each concept is assigned a unique identifier called a Concept Unique Identifier (CUI). CUIs are alphanumeric codes that remain stable across different versions of UMLS, serving as a means to identify and link concepts across multiple vocabularies and ontologies. For example, the CUI C0018787 represents the concept of “Heart” in UMLS. In this representation, text documents were encoded as vectors where each element corresponded to a unique CUI. The presence of a CUI was indicated by a non-zero value, which also encoded contextual information such as assertion status (e.g., positive or negative). This approach combines elements of one-hot encoding with additional semantic information, allowing for a more nuanced representation of the document’s content.

#### Machine Learning Training, Optimization, and Evaluation Metrics

This study employed a rigorous methodology to train, optimize, and compare three machine learning algorithms: RF, XGBoost, and KNN. We utilized a weakly supervised learning approach, training models on silver-labeled documents to learn discriminative features for identifying clinically relevant notes without extensive manual annotation. While not perfect, this silver-labeled dataset provided a valuable starting point for model training.

The optimization process starts with hyperparameter tuning through grid search, systematically examining key parameters for each model. For RF, we considered the number of trees, maximum depth, and minimum samples split. XGBoost optimization focused on learning rate, maximum depth, subsample, and number of boosting rounds. For KNN, we tuned the number of neighbors and distance metric. Five-fold cross-validation assessed each hyperparameter combination, with the F1-score serving as the primary metric due to its capacity to balance precision and recall.

To refine our approach, we leveraged a small set of clinician-annotated evaluation data. This allowed for better fine-tuning of the models to distinguish document relevance, balancing the primary aim of maximizing recall (to ensure identification of as many truly relevant documents as possible) with maintaining precision (to avoid excessive misclassification of irrelevant documents). We proceeded to fine-tune the models by adjusting classification thresholds. Probability predictions were generated on a validation dataset, with thresholds ranging from 0.0 to 1.0 in 0.1 increments.

Precision, recall, and F1-score were calculated at each threshold, facilitating the identification of an optimal threshold that maximized the F1-score while maintaining an appropriate balance between precision and recall. The final evaluation was conducted on a held-out test set, focusing primarily on key metrics: recall (sensitivity), precision (positive predictive value), specificity, and F1-score for the relevant class. Recall measures the fraction of total relevant documents correctly classified, while precision assesses the accuracy in identifying only relevant documents, minimizing false positives^44^.

Specificity evaluates the system’s ability to correctly identify non-relevant documents, providing a comprehensive picture of classification performance. The F1-score, as the harmonic mean of precision and recall, offers a combined measure of classification accuracy^44^. Detailed performance metrics at various classification thresholds for the best-performing model are provided in the appendix, offering deeper insights into model behavior across diverse scenarios.

### 3. Experiment Design

#### Baseline

The baseline classification for COPD-related documents uses expert-defined heuristics based on ICD codes and the temporal relevance of notes. These heuristics were evaluated against a manually annotated gold standard test dataset, which included examples from each category (COPD-coded, non-COPD-coded, and pre-COPD temporal). This evaluation serves two purposes: first, to assess the effectiveness of these simple rules in identifying COPD-relevant notes compared to clinician annotations, and second, to establish a baseline for comparison with machine learning approaches. The subsequent machine learning models are trained on the heuristically labeled data, optimized using a small annotated dataset, and then tested on the same gold standard test data. This process determines if machine learning can optimize the classification process while requiring only a small annotated dataset for fine-tuning.

#### Experiment Setup

We experimented with three classifiers - RF, XGBoost, and KNN - using BoW representations, as these models are known to effectively handle various data types ^45,46^. Following the same standardized procedure, we also experimented with the machine learning models using other text representations - TF-IDF, lightweight document embeddings, and compression-based features.

#### Hyperparameter Tuning

Hyperparameter tuning was conducted for RF, XGBoost, and KNN classifiers to optimize their performance on the text classification task. For RF, we explored the number of trees (100-500), maximum depth (10-30, None), minimum samples for split and leaf nodes (2-10 and 1-4 respectively), feature selection strategies for node splitting, and class weight options. XGBoost tuning focused on learning rate (0.01-0.3), number of estimators (100-1000), maximum depth (3-10), minimum child weight (1-6), subsampling ratio (0.5-1.0), and column sampling by tree (0.5-1.0). For KNN, we varied the number of neighbors (1-20), weighting function (uniform, distance), distance metrics (Euclidean, Manhattan, Minkowski), and leaf size (10-50).

GridSearchCV with 5-fold cross-validation was employed for all models, using multiple scoring metrics (F1-score, precision, recall, and specificity) to evaluate performance. This approach allowed for the selection of optimal hyperparameter combinations that balance model complexity, computational efficiency, and generalization performance, tailored to the specific challenges of text classification

#### Threshold Analysis

In our case, optimizing recall is crucial, particularly because the cost of false negatives is high. While classification models often default to a 0.5 threshold for converting probability predictions into class labels, this threshold may not be ideal for our needs. Our threshold analysis evaluates model performance across a range of thresholds, from 0.0 to 1.0, in increments of 0.1. At each threshold, we calculate precision, recall, specificity, and the F1-score. This analysis allows us to fine-tune the model’s decision boundary to maximize recall while maintaining excellent precision, ensuring that we identify as many relevant cases as possible without significantly increasing the number of false positives.

For all experiments, models were optimized on a separate validation set to find the optimal hyperparameters that maximize recall, as accurately identifying all relevant documents is crucial.

Instead of relying solely on the default threshold, we iteratively adjusted the recall threshold to find the optimal cutoff for each model. The optimized parameters based on this evaluation were then used in the final models and applied to the held-out test set. By adhering to this consistent methodology, we were able to fairly assess the performance of different classifiers and text representations.

## Results

### Baseline Performance

The defined ICD-based heuristics are evaluated against manually annotated gold standard test data. Table 1 shows the baseline model’s performance in classifying COPD-related documents. Figure 2 shows the baseline confusion matrix on the test data. The heuristics correctly identified 50 irrelevant and 57 relevant documents while misclassifying 24 irrelevant and 92 relevant documents.

**Table 1.** shows the baseline model’s performance in classifying COPD-related documents.

| Baseline | Precision (PPV) | Recall(sensitivity) | Specificity | F1-score (95%CI) |
| --- | --- | --- | --- | --- |
|  | 0.70 | 0.38 | 0.68 | 0.50 (0.43 - 0.56) |

**Figure 2.**
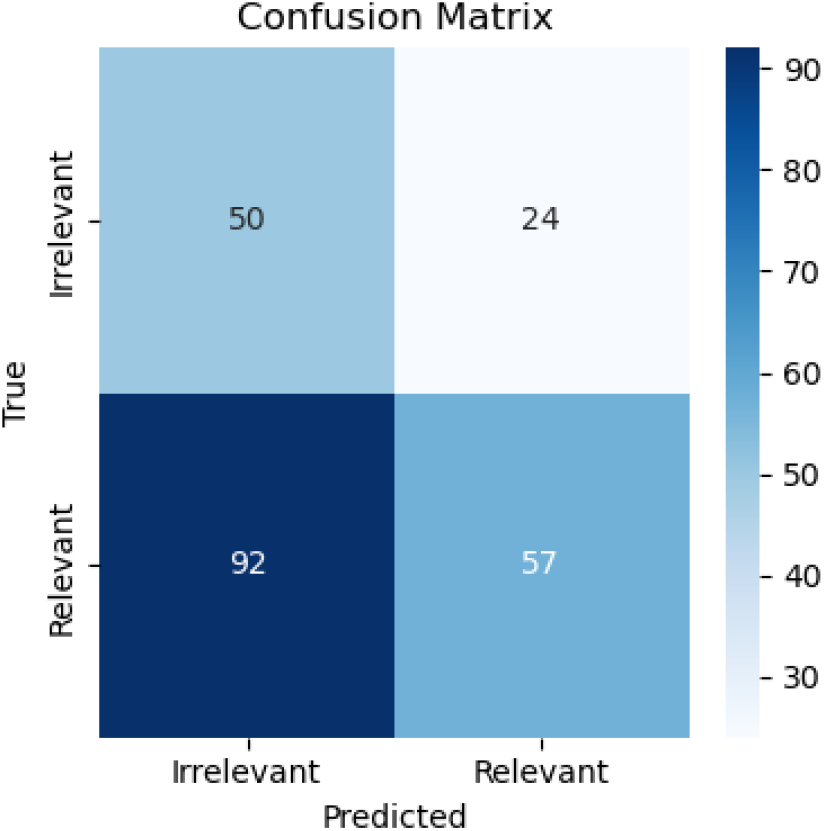
Baseline confusion matric on the test data

### Machine learning framework performance

The results validate the effectiveness of our framework, which uses various text representation techniques and machine learning models to eliminate irrelevant documents and prioritize information in relevant ones. We found that combining both negative cases (non-COPD-coded and pre-COPD temporal) consistently produced the best model across all representation techniques. The supplementary Tables (Tables S1 to S5) provide comprehensive results for all combinations, but here, we focus on the results obtained by training the model on the combined dataset using both negative heuristics.

Table 2 presents the performance comparison of different classification models using various text representation techniques on the COPD dataset.

**Table 2.** Performance Comparison of Classification Models on COPD Dataset using various Representation.

| Representation | Model | Precision (PPV) | Recall (Sensitivity) | Specificity | F1-score (95% CI) |
| --- | --- | --- | --- | --- | --- |
| BoW Representation | RF | 0.77 | 0.71 | 0.57 | 0.74 (0.7-0.78) |
|  | XGBoost | 0.75 | 0.67 | 0.54 | 0.71 (0.65-0.76) |
|  | KNN | 0.71 | 0.47 | 0.62 | 0.57 (0.53-0.62) |
| TF-IDF Representation | RF | 0.76 | 0.69 | 0.57 | 0.73 (0.68-0.78) |
|  | XGBoost | 0.71 | 0.64 | 0.46 | 0.67 (0.61-0.71) |
|  | KNN | 0.80 | 0.37 | 0.81 | 0.51(0.46-0.55) |
| Lightweight Document Embedding Representation | RF | 0.75 | 0.89 | 0.39 | 0.81 (0.76-0.87) |
|  | XGBoost | 0.73 | 0.74 | 0.45 | 0.74 (0.71-0.77) |
|  | KNN | 0.89 | 0.32 | 0.92 | 0.47 (0.43-0.52) |
| Compression-based Representation | RF | 0.78 | 0.45 | 0.74 | 0.57 (0.52-0.6) |
|  | XGBoost | 0.78 | 0.45 | 0.74 | 0.57 (0.54-0.62) |
|  | KNN | 0.75 | 0.64 | 0.57 | 0.69 (0.63-0.74) |
| UMLS Text Representation | RF | 0.74 | 0.55 | 0.64 | 0.63 (0.58-0.69) |
|  | XGBoost | 0.76 | 0.56 | 0.46 | 0.65 (0.6-0.68) |
|  | KNN | 0.86 | 0.30 | 0.86 | 0.44 (0.38-0.51) |

Table 2 presents a comparative analysis of various ML models (RF, XGBoost, and KNN) across different document representations. The RF model consistently demonstrates superior performance with a balanced precision, recall, and F1-score for the relevant class when using BoW representation. In the TF-IDF representation, KNN shows high precision but lower recall, while RF provides a more balanced performance. With lightweight document embeddings, RF achieves high recall but lower precision for the irrelevant class, indicating a trade-off. For compression-based representations, RF and XGBoost perform comparably, with KNN achieving a higher F1 score. Lastly, in the UMLS representation, XGBoost marginally outperforms RF in precision and F1-score, while KNN shows high precision but lower recall.

#### Results summary

Lightweight document embedding combined with the RF model was the most effective method for extracting COPD-relevant documents. This approach achieves an F1-score of 0.81 (0.76-0.87) for the Relevant class, outperforming other representation techniques examined in this study.

### Best Performing Model Comparison and Threshold Analysis

The baseline model confusion matrix (Figure 2) correctly identified 50 irrelevant and 57 relevant documents, with 24 irrelevant and 92 relevant misclassified. The best-performing NLP pipeline confusion matrix (Figure 3) correctly identified 29 irrelevant and 128 relevant documents, with 47 irrelevant and 23 relevant misclassified. The best-performing model significantly improves relevant document identification (128 vs. 57) and reduces relevant misclassification (21 vs. 92).

**Figure 3.**
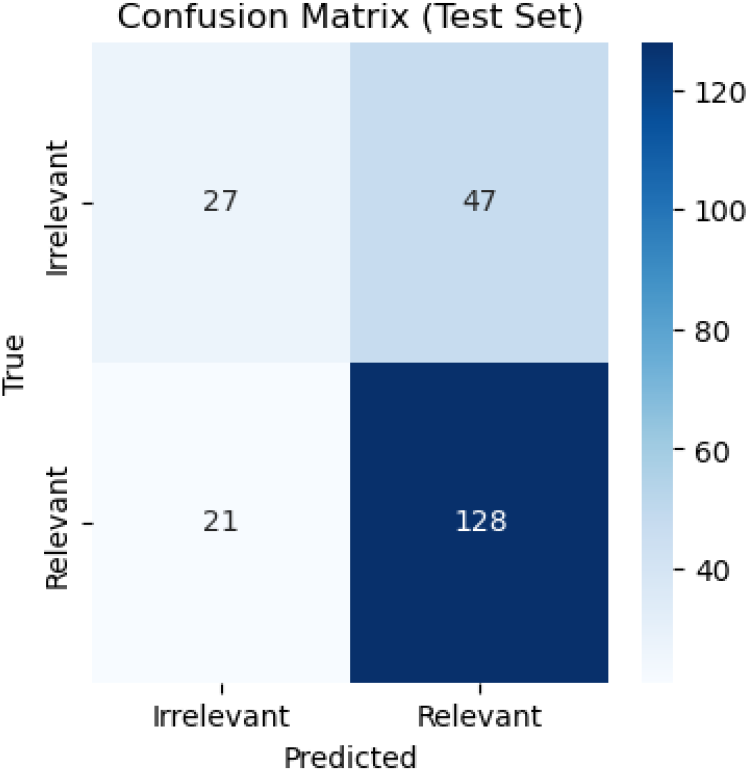
best performing NLP pipeline confusion matrix

Although the best-performing NLP pipeline has a higher irrelevant misclassification rate (as highlighted by low specificity), this trade-off is acceptable given the optimization for sensitivity, which was the primary goal. The detailed results for the best-performing classifier at various cutoff points are presented in Appendix Figure S1. The figure illustrates the relationship between precision, sensitivity, and specificity across different thresholds for the classification model.

## Discussion

Our study demonstrates the effectiveness of lightweight machine learning models, particularly a RF classifier with lightweight document embeddings, in extracting COPD-relevant documents from EHRs. This approach significantly outperformed baseline heuristic methods, achieving high recall (0.89) and F1-score (0.81, CI: 0.76–0.87) for relevant documents. These findings represent a significant advancement in efficient EHR processing, offering a scalable solution for filtering large volumes of clinical documents in resource-constrained environments. By improving the identification of relevant information, this approach has the potential to enhance the performance of clinical decision support systems and optimize resource utilization in healthcare applications.

### Machine learning Optimization Over Heuristic Approaches

Traditional heuristic approaches, such as relying on ICD codes for document filtering, have proven inadequate in accurately capturing relevant clinical information. Our findings confirm that a substantial proportion of documents with COPD ICD codes lack significant relevant information (as illustrated in Figure 2 above), highlighting the limitations of such methods. In contrast, our machine learning framework effectively distinguishes relevant from irrelevant documents (as illustrated in Figure 3 above), thereby improving the sensitivity and identification of relevant documents. Machine learning models outperform baseline approaches by optimizing small annotations, learning deeper patterns, adapting flexibly, generalizing better from imperfect examples, handling noisy data robustly, and using advanced feature extraction techniques ^16,47^.

### Improvement and Effectiveness of the Model

A primary contribution of our work is demonstrating how lightweight models, such as a RF classifier paired with lightweight document embeddings, can achieve high performance with minimal annotation data. By employing weak supervision, we generated “silver standard” labels from domain-expert heuristics, significantly reducing the need for extensive manual annotation. This method not only streamlined the training process but also enhanced the model’s scalability across different clinical domains.

Our methodology uniquely integrates a large corpus of weakly supervised “silver standard” labels for initial training with a small set of high-fidelity “gold standard” labels for hyperparameter optimization. This approach not only streamlined the training process but also enhanced model performance and generalizability across diverse clinical domains.

As shown in Table 2, among the evaluated text representation techniques, lightweight document embedding coupled with the RF classifier exhibited superior performance in classifying relevant COPD documents, achieving a recall of 0.89 and an F1-score of 0.81 (95% CI: 0.76-0.87) for the relevant class. This performance significantly surpassed the baseline model’s F1-score of 0.50 (95% CI: 0.43-0.56), underscoring the efficacy of our approach compared to conventional heuristic methods.

The incorporation of a limited set of high-fidelity labels for hyperparameter tuning resulted in a substantial improvement in model performance. The model-agnostic nature of this approach renders it applicable to any classifier requiring hyperparameter optimization.

Additionally, the results underscore the critical role of text representation in model performance. While both RF and XGBoost proved to be robust classifiers, their effectiveness was notably influenced by the choice of text representation technique. For instance, RF’s F1-score for the relevant class ranged from 0.57 (95% CI: 0.52-0.62) with compression-based representation to 0.81 (95% CI: 0.76-0.87) with lightweight document embedding.

### Limitations and Future Directions

While our study demonstrates the effectiveness of lightweight document embedding and machine learning models in extracting relevant COPD documents, a number of limitations require consideration. First, the dataset used in this study was specific to COPD, and the generalizability of our findings to other clinical domains requires further investigation. Future research should explore the applicability of our framework to a broader range of clinical conditions and document types.

Additionally, while our study focused on the binary classification of documents as relevant or irrelevant, future work could explore a more granular categorization of clinical documents based on their specific content or purpose. This multi-class classification approach could provide even more targeted information for clinical decision support systems.

## Conclusion

This study presents a novel framework for extracting COPD-relevant clinical documents using lightweight document embedding and machine learning models. Our approach effectively identifies relevant documents while minimizing irrelevant ones, enhancing the quality of information for clinical decision-support systems and improving patient outcomes. Future research should explore the generalizability of our findings to other chronic conditions and integrate this model into predictive analytics for greater efficiency and effectiveness. This framework can be adopted across diverse healthcare domains, contributing to improved patient care and clinical outcomes.

## Data Availability

All data produced in the present study are available upon reasonable request to the authors

## Supplementary

**Table S1.**
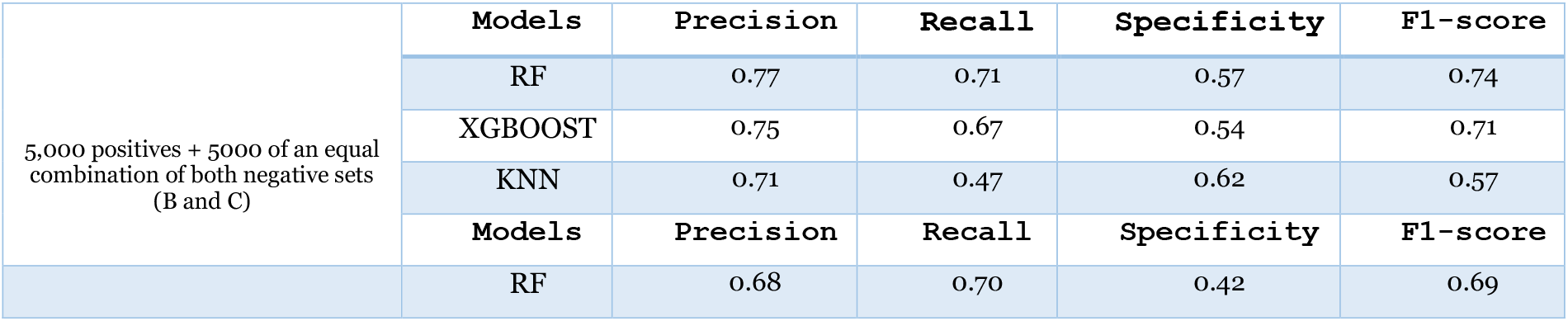
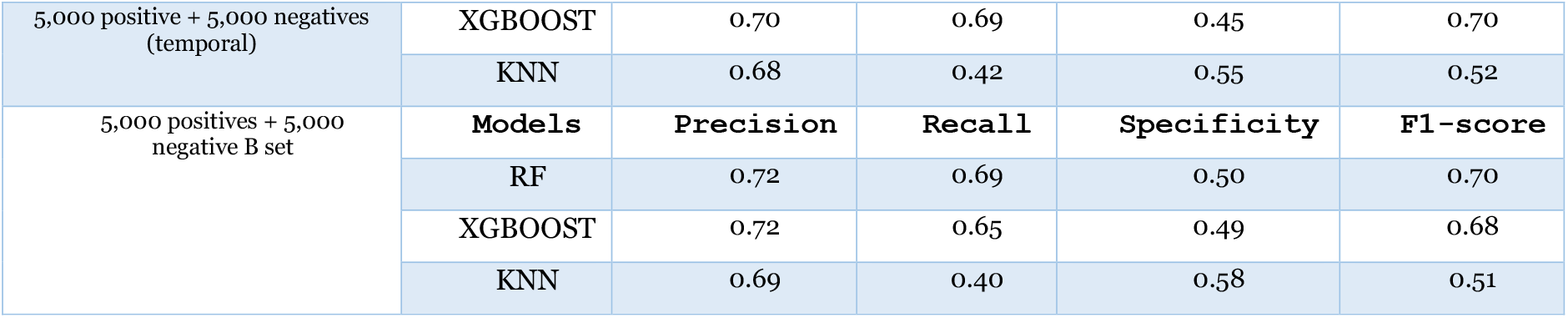
Performance Comparison of Classification Models on COPD Dataset using BoW Representation.

**Table S2.** Performance Comparison of Classification Models on COPD Dataset using TF-IDF Representations.

|  |  |  |  |  |  |
| --- | --- | --- | --- | --- | --- |
| 5,000 positives + 5000 of an equal combination of both negative sets (B and C) | **Models** | **Precision** | **Recall** | **Specificity** | **F1-score** |
|  | RF | 0.76 | 0.69 | 0.57 | 0.73 |
|  | XGBOOST | 0.71 | 0.64 | 0.46 | 0.67 |
|  | KNN | 0.80 | 0.37 | 0.81 | 0.50 |
|  | **Models** | **Precision** | **Recall** | **Specificity** | **F1-score** |
| 5,000 positives + 5,000 negative B set | RF | 0.71 | 0.60 | 0.45 | 0.60 |
|  | XGBOOST | 0.69 | 0.62 | 0.42 | 0.65 |
|  | KNN | 0.76 | 0.36 | 0.63 | 0.49 |
| 5,000 positives + 5,000 negative C set | **Models** | **Precision** | **Recall** | **Specificity** | **F1-score** |
|  | RF | 0.76 | 0.69 | 0.57 | 0.72 |
|  | XGBOOST | 0.71 | 0.63 | 0.42 | 0.66 |
|  |  | 0.39 | 0.42 |  | 0.40 |
|  | KNN | 0.78 | 0.33 | 0.68 | 0.47 |

**Table S3.**
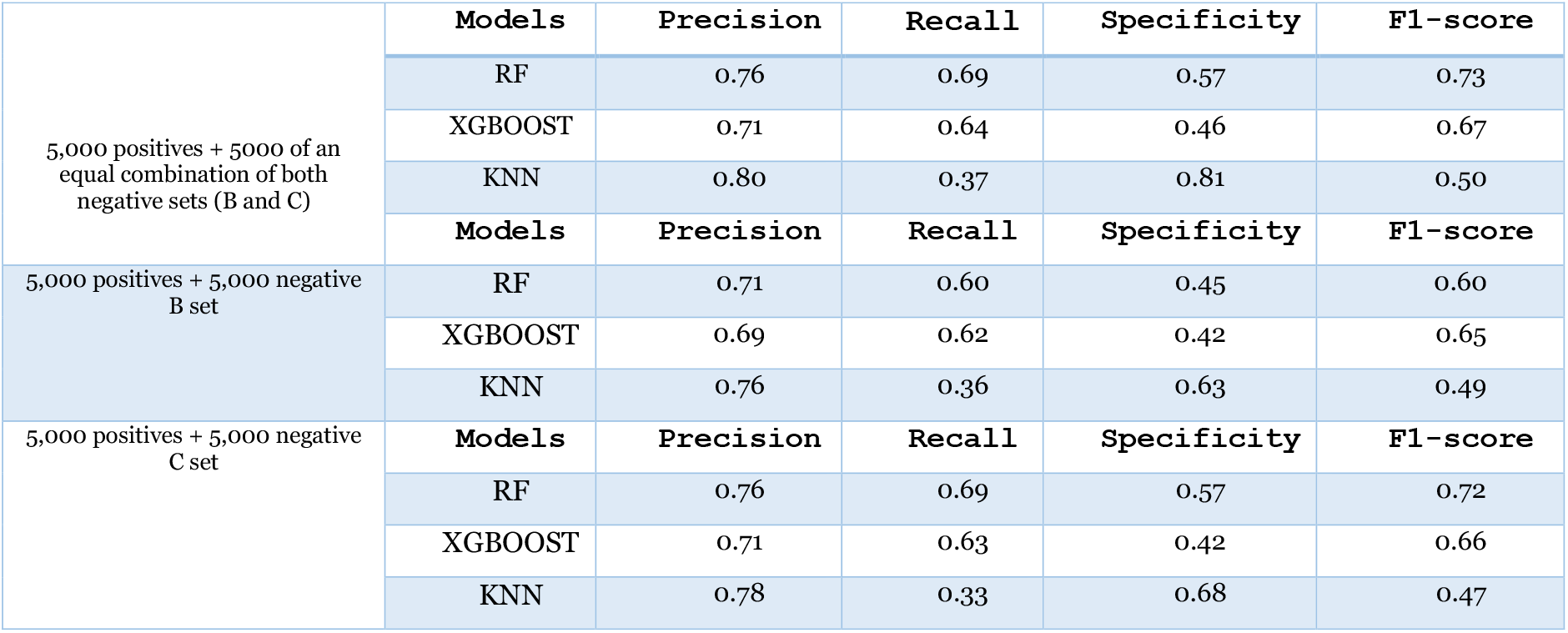
Performance Comparison of Classification Models on COPD using Lightweight Document Embedding Representation.

|  |  |  |  |  |  |
| --- | --- | --- | --- | --- | --- |
| 5,000 positives + 5000 of an equal combination of both negative sets (B and C) | **Models** | **Precision** | **Recall** | **Specificity** | **F1-score** |
|  | RF | 0.76 | 0.69 | 0.57 | 0.73 |
|  | XGBOOST | 0.71 | 0.64 | 0.46 | 0.67 |
|  | KNN | 0.80 | 0.37 | 0.81 | 0.50 |
|  | **Models** | **Precision** | **Recall** | **Specificity** | **F1-score** |
| 5,000 positives + 5,000 negative B set | RF | 0.71 | 0.60 | 0.45 | 0.60 |
|  | XGBOOST | 0.69 | 0.62 | 0.42 | 0.65 |
|  | KNN | 0.76 | 0.36 | 0.63 | 0.49 |
| 5,000 positives + 5,000 negative C set | **Models** | **Precision** | **Recall** | **Specificity** | **F1-score** |
|  | RF | 0.76 | 0.69 | 0.57 | 0.72 |
|  | XGBOOST | 0.71 | 0.63 | 0.42 | 0.66 |
|  | KNN | 0.78 | 0.33 | 0.68 | 0.47 |

**Table S4.** Performance Comparison of Classification Models on COPD using Compression-based Representation.

|  |  |  |  |  |  |
| --- | --- | --- | --- | --- | --- |
|  | **Models** | **Precision** | **Recall** | **Specificity** | **F1-score** |
|  | RF | 0.78 | 0.45 | 0.74 | 0.57 |
| 5,000 positives + 5000 of an equal combination of both negative sets (B and C) | XGBOOST | 0.78 | 0.45 | 0.74 | 0.57 |
|  | KNN | 0.75 | 0.64 | 0.57 | 0.69 |
|  | **Models** | **Precision** | **Recall** | **Specificity** | **F1-score** |
| 5,000 positives + 5,000 negative B set | RF | 0.62 | 0.41 | 0.63 | 0.49 |
|  | XGBOOST | 0.71 | 0.47 | 0.71 | 0.57 |
|  | KNN | 0.68 | 0.57 | 0.52 | 0.62 |
| 5,000 positives + 5,000 negative C set | **Models** | **Precision** | **Recall** | **Specificity** | **F1-score** |
|  | RF | 0.76 | 0.42 | 0.71 | 0.54 |
|  | XGBOOST | 0.73 | 0.28 | 0.72 | 0.41 |
|  | KNN | 0.69 | 0.60 | 0.5 | 0.64 |

**Table S5.** Performance Comparison of Classification Models on COPD using UMLS Representation.

|  |  |  |  |  |  |
| --- | --- | --- | --- | --- | --- |
| 5,000 positives + 5000 of an equal combination of both negative sets (B and C) | **Models** | **Precision** | **Recall** | **Specificity** | **F1-score** |
|  | **RF** | 0.74 | 0.55 | 0.64 | 0.63 |
|  | XGBOOST | 0.76 | 0.56 | 0.46 | 0.65 |
|  | KNN | 0.86 | 0.30 | 0.86 | 0.44 |
|  | **Models** | **Precision** | **Recall** | **Specificity** | **F1-score** |
| 5,000 positives + 5,000 negative B set | **RF** | 0.69 | 0.51 | 0.62 | 0.59 |
|  | XGBOOST | 0.70 | 0.51 | 0.42 | 0.59 |
|  | KNN | 0.80 | 0.25 | 0.76 | 0.38 |
| 5,000 positives + 5,000 negative C set | **Models** | **Precision** | **Recall** | **Specificity** | **F1-score** |
|  | **RF** | 0.64 | 0.45 | 0.54 | 0.53 |
|  | XGBOOST | 0.72 | 0.46 | 0.42 | 0.56 |
|  | KNN | 0.82 | 0.25 | 0.72 | 0.38 |

**Figure S1:**
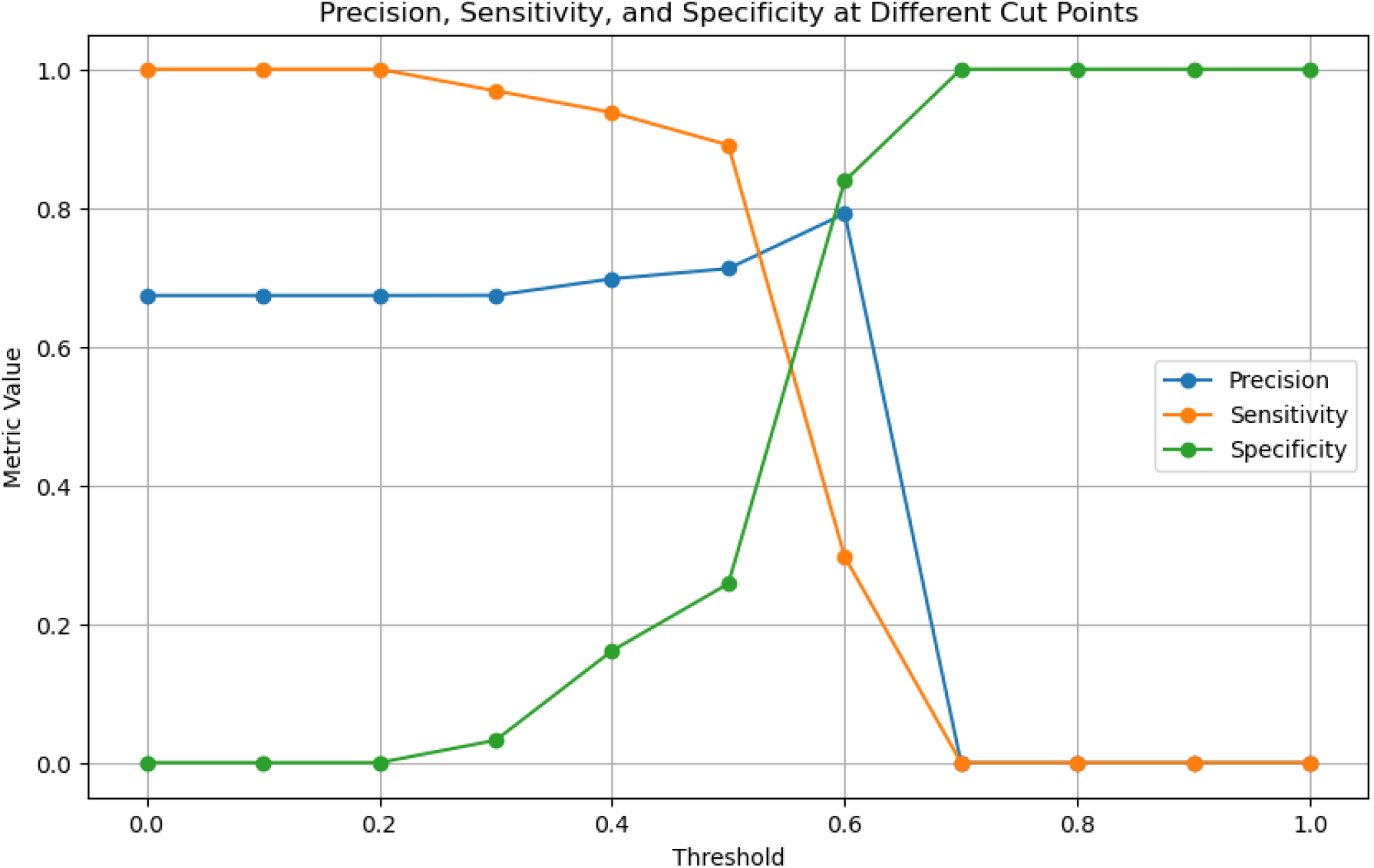
Variation of Precision, Sensitivity, and Specificity Across Different Thresholds

